# Symptom-level genetic modelling identifies novel risk loci and unravels the shared genetic architecture of anxiety and depression

**DOI:** 10.1101/2020.04.08.20057653

**Authors:** Jackson G. Thorp, Adrian I. Campos, Andrew D. Grotzinger, Zachary Gerring, Jiyuan An, Jue-Sheng Ong, Wei Wang, 23andMe Research Team, Suyash Shringarpure, Enda M. Byrne, Stuart MacGregor, Nicholas G. Martin, Sarah E. Medland, Christel M. Middeldorp, Eske M. Derks

## Abstract

Depression and anxiety are highly prevalent and comorbid psychiatric traits that cause considerable burden worldwide. Previous studies have revealed substantial genetic overlap between depression, anxiety, and a closely related personality trait – neuroticism. Here, we use factor analysis and genomic structural equation modelling (Genomic SEM) to investigate the genetic factor structure underlying 28 items assessing depression, anxiety and neuroticism. Symptoms of depression and anxiety loaded on two distinct, although genetically correlated factors, while neuroticism items were partitioned between them. We leveraged this factor structure to conduct multivariate genome-wide association analyses on latent factors of anxiety symptoms and depressive symptoms, using data from over 400,000 individuals in the UK Biobank. We identified 89 independent variants for the depressive factor (61 genomic loci, 29 novel) and 102 independent variants for the anxiety factor (73 loci, 71 novels). Of these variants, 72% and 78%, respectively, replicated in an independent 23andMe cohort of ∼1.9 million individuals with self-reported diagnosis of depression (634,037 cases) and anxiety (624,615 cases). A pairwise GWAS analysis revealed substantial genetic overlap between anxiety and depression but also showed trait-specific genetic influences; e.g. genomic regions specific to depressive symptoms were associated with hypertriglyceridemia, while regions specific to anxiety symptoms were linked to blood pressure phenotypes. The substantial genetic overlap between the two traits was further evidenced by a lack of trait-specificity in polygenic prediction of depressive and anxiety symptoms. Our results provide novel insight into the genetic architecture of depression and anxiety and comorbidity between them.

---

Depression and anxiety are the two most prevalent psychiatric disorders and cause substantial disease burden, accounting for over 10% of years lived with disability worldwide (Vigo, Thornicroft, and Atun 2016; World Health Organization 2017). They are highly comorbid; around three quarters of people with an anxiety disorder also meet diagnostic criteria for major depressive disorder (Lamers et al. 2011). Genetic factors play a substantial role in liability to these disorders, with heritability estimates between 30 to 40 percent for both depression and anxiety (Hettema, Neale, and Kendler 2001; Sullivan, Neale, and Kendler 2000). Twin and family studies suggest that their comorbidity is largely explained by shared genetic risk factors (Middeldorp et al. 2005).

Neuroticism, characterised as the tendency to experience emotional negativity, such as mood swings, sadness, and worry (McCrae and Costa 1985; Eysenck and Eysenck 1985), is a shared risk factor for depression and anxiety (Kotov et al. 2010; Gray and McNaughton 2000; Ormel et al. 2013; Zinbarg et al. 2016). Genetic factors explain around ∼40% of variation in neuroticism (Vukasović and Bratko 2015), and these factors largely overlap with those that affect depression and anxiety (Hettema, Prescott, and Kendler 2004; Jardine, Martin, and Henderson 1984; Fanous et al. 2002; Hettema et al. 2006; Middeldorp et al. 2005). Recent molecular genetic studies have uncovered extensive pleiotropy between these three traits (Purves et al. 2019; Meier et al. 2019; Wray et al. 2018), but little is known about their genetic overlap at a symptom-based level. Here, we investigate the genetic relationship between individual symptoms of anxiety, depression and neuroticism, in order to elucidate their genetic overlap and gain insight into the biological mechanisms underlying comorbidity between anxiety and depression.

Genome-wide association studies (GWAS) have accelerated our progress in unravelling the genetic architecture of these psychiatric traits. The general observation is that complex traits are influenced by large numbers of genetic variants with small individual effect sizes (i.e. high polygenicity) and consequently require very large sample sizes in order to detect them. Recent GWAS have identified over 100 independent and robustly associated single nucleotide polymorphisms (SNPs) for depression and neuroticism (Howard et al. 2019; Wray et al. 2018; Nagel, Jansen, et al. 2018; Luciano et al. 2018). In comparison, genetic studies of anxiety are still underpowered, with the largest studies to date having identified 5 genetic risk loci for lifetime anxiety disorder (Purves et al. 2019) and 6 loci for anxiety symptoms (Levey et al. 2020). While the full-scope of risk conferring genetic loci remains to be discovered for these traits, bivariate genomic methods (e.g., LD Score Regression; Bulik-Sullivan et al. 2015) have been used to obtain estimates of overall levels of genetic overlap. Consistent with family-based results, large SNP-based genetic correlations have been reported across depression, anxiety, and neuroticism (*r*_g_ > 0.70) (Wray et al. 2018; Nagel, Jansen, et al. 2018). Moreover, pairwise comparison of genomic loci implicated in neuroticism and major depression found a substantial portion (∼70%) of regions associated with major depression are also associated with neuroticism (Adams et al. 2019).

The extensive genetic overlap between depression, anxiety, and neuroticism may partly reflect overlap in item content and diagnostic criteria used to measure these traits (Ormel, Riese, and Rosmalen 2012). Numerous scales of neuroticism include “sub-scales” or “facets” of both depression and anxiety (e.g. NEO Personality Inventory-Revised (NEO-PI-R); California Psychological Inventory (CPI) Big Five), and many items within these scales closely resemble symptom measures of both depression and anxiety. For example, NEO-PI-R neuroticism items “*Sometimes I feel completely worthless*” and *“I have sometimes experienced a deep sense of guilt or sinfulness”* are very similar to the DSM-5 major depression symptom “*Feelings of worthlessness or excessive or inappropriate guilt”*. Neuroticism is therefore not operationally distinct, implying that by studying its underlying components one could gain valuable insight into symptoms of anxiety and depression and their genetic influences. Indeed, hierarchical clustering of individual neuroticism items in the Short-form Eysenck Personality Questionnaire-Revised (EPQR-S) (Eysenck, Eysenck, and Barrett 1985) revealed two genetic item clusters, termed ‘depressed affect’ and ‘worry’, displaying stronger genetic overlap with depression and anxiety, respectively (Nagel, Watanabe, et al. 2018).

Item or symptom-level genetic analyses allow investigation of the underlying genetic structure of a trait and have proven useful in disentangling genetic and phenotypic heterogeneity of neuroticism and depression (Nagel, Watanabe, et al. 2018) (Thorp et al. 2019). In the present study, we extend this approach across multiple traits and investigate the genetic factor structure underlying 28 symptoms of depression, anxiety, and neuroticism. We apply genomic structural equation modelling (Genomic SEM) (Grotzinger et al. 2019), a recently developed multivariate method which enables estimation of the joint genetic architecture of multiple complex traits based on summary statistics from GWAS. This allows genetic subtypes or combinations of genetically similar symptoms to be identified, leading to increased statistical power for the discovery of genetic loci and improved understanding of the comorbidity and genetic overlap across traits. We sought to answer three questions: (1) how do items used to measure neuroticism genetically relate to symptoms of depression and anxiety; (2) can we leverage genetic overlap with neuroticism to boost power for the discovery of genetic risk loci for anxiety and depressive symptoms; and (3) can we identify genetic loci that differentiate anxiety and depressive symptoms? First, we model the genetic factor structure across the three traits using item-level questionnaire data from the UK Biobank (anxiety and depressive symptoms, N = ∼135,000; neuroticism, N = ∼400,000). Then, we leverage this factor structure to identify genetic loci for latent factors of depressive symptoms and anxiety symptoms using Genomic SEM. Finally, we identify genomic regions that are unique to or shared by depressive and anxiety symptoms to gain insight into the genetic architecture of these traits and the comorbidity between them.

## Results

### Factor analysis of symptoms of depression, anxiety, and neuroticism

We explored genetic overlap between anxiety symptoms, depressive symptoms, and neuroticism by modelling the genetic factor structure of items used to measure these traits. Item-level genome-wide association analyses were conducted individually on each of 28 items of neuroticism (12 items; EPQR-S), anxiety (7 items; GAD-7), and depression (9 items; PHQ-9), in ∼135,000 UKB participants (see Supplementary table 1 for item-specific sample sizes). LD Score regression was used to calculate genetic correlations between all item pairs and an exploratory factor analysis (EFA) was then conducted on this genetic correlation matrix. A minimum average partial (MAP) test suggested the optimal number of factors to extract is three (consistent with the eigenvalue-greater-than-one rule; see Supplementary Table 2).

Factor loadings of the 3-factor model are presented in Figure 1 (and Supplementary Table 3). Depression items had high loadings (> 0.4) on genetic factor 1, anxiety items had high loadings on genetic factor 2 (except for the item ‘irritability’, which loaded onto factor 1), and neuroticism items loaded either highly on factor 1 (5 items) or factor 2 (7 items). Genetic factor 3 is characterised by relatively low loadings, which are positive for items of depression and anxiety, and negative for items of neuroticism. The items with the highest loadings on factor 3 are mostly somatic symptoms; therefore this factor may contain variance that separates a psychosomatic facet of depression and anxiety from neuroticism. Factor 3 is largely underpowered for further genetic analysis; we therefore restrict subsequent analyses to factor 1 and factor 2. In this paper, we refer to these two genetic latent factors as DEP (depressive symptoms) and ANX (anxiety symptoms).

**Figure 1:**
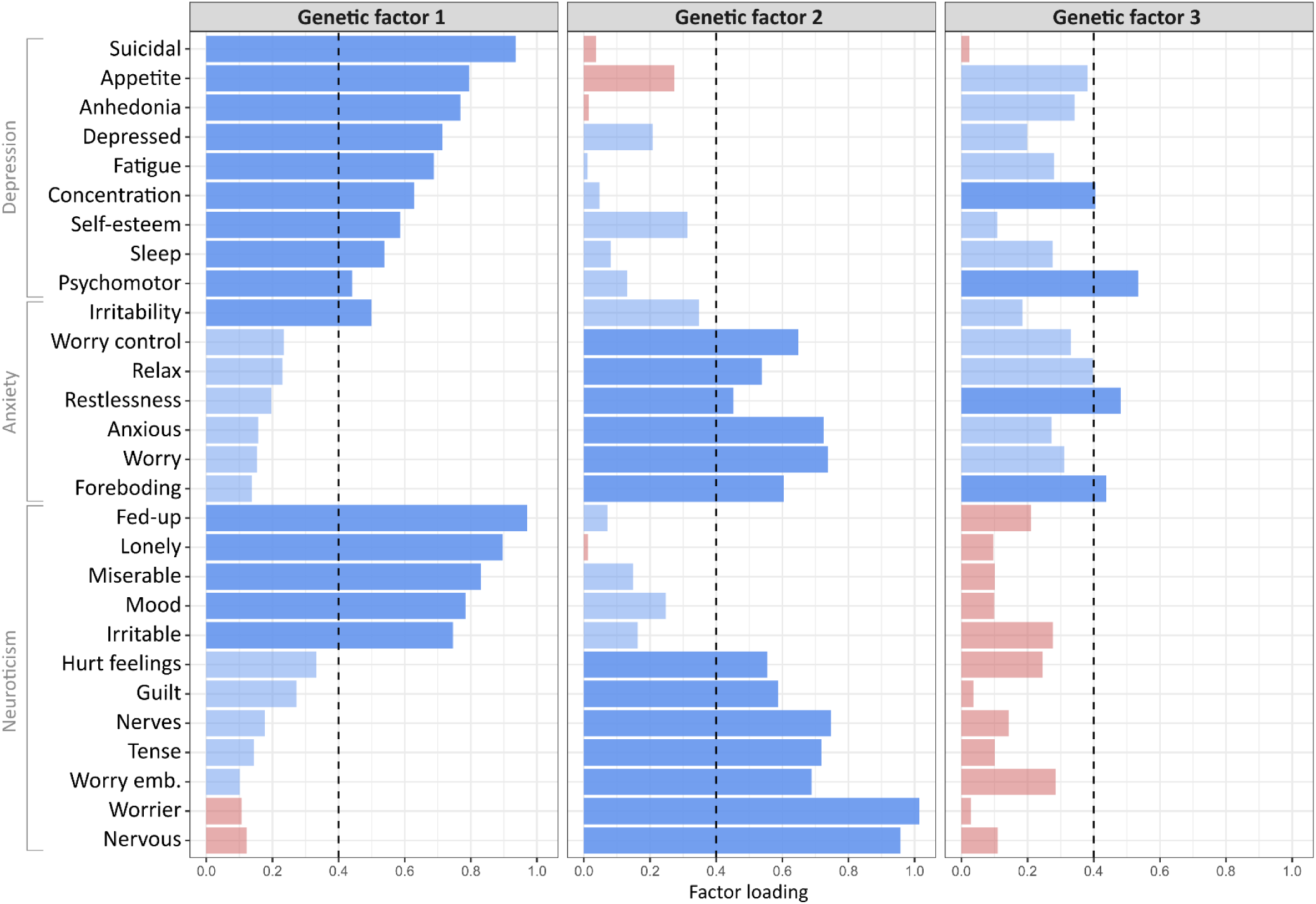
Standardised factor loadings from a genetic exploratory factor analysis of 28 items of depression, anxiety and neuroticism. Note: Positive loadings are indicated in blue and negative loadings in red. Items with a standardised loading less than 0.4 are shown as transparent.

We then submitted this factor structure model to Genomic SEM (retaining standardised loadings > 0.4) to assess its fit to the data (taking into account uncertainty in covariance estimates). The model provided adequate fit to the data (CFI = 0.890; SRMR = 0.087; factor loadings in Supplementary Table 4). As a first validation of the DEP and ANX latent factors that included the neuroticism items, we calculated genetic correlations between the latent factors and sum-scores of depressive symptoms (PHQ-9; N = 135,149) and anxiety symptoms (GAD-7; N = 135,747). The genetic correlations between the PHQ-9 sum-score and the DEP latent factor (rg = 0.94, 95% CI [0.87,1.01]), and the GAD-7 sum-score and ANX latent factor (rg = 0.93, 95% CI [0.86,1.00]) were not significantly different from one, suggesting the DEP and ANX latent factors are good proxies for anxiety and depressive symptoms. The genetic correlation between the DEP and ANX factors was moderately high (rg = 0.80, 95% CI [0.77,0.83]) and was similar to the correlation between the PHQ-9 and GAD-7 sum-scores (rg = 0.83, 95% CI [0.72,0.95]).

### Multivariate GWAS of anxiety and depressive symptoms

Having identified the genetic latent factor structure within the UKB sample, our next step was to leverage this structure to identify genomic risk loci for the DEP and ANX latent factors. To maximise power in the multivariate GWAS, we expanded the neuroticism items to the full UKB set (i.e. an additional ∼270k individuals who completed the neuroticism questionnaire but not the depressive or anxiety symptoms questionnaires were included; each neuroticism item N = ∼400k). We examined whether this changed the factor structure by fitting the EFA-derived model to the genetic covariance matrix of the full UKB set. The EFA-derived factor structure retained adequate fit to the data (CFI = 0.893; SRMR = 0.088), while standardised factor loadings were concordant with loadings before expanding the sample size of neuroticism (r = 0.95; factor loadings in Supplementary Table 5). The genetic correlation between the DEP and ANX factors remained the same (rg = 0.79, 95% CI [0.77,0.81]).

Multivariate GWAS were conducted by estimating the effects of individual SNPs on the DEP and ANX latent factors using Genomic SEM. The GWAS of the DEP factor identified 7,677 genome-wide significant SNPs (*P* < 5 × 10^−8^), tagging 89 independent SNPs in 62 genomic risk loci (see Figure 2a and Supplementary Table 6). Of these loci, 33 overlap with a previous GWAS of depressive symptoms or major depression (Wray et al. 2018; Nagel, Jansen, et al. 2018; Howard et al. 2019; Okbay et al. 2016; Howard et al. 2018; Hyde et al. 2016; Turley et al. 2018; Baselmans et al. 2019), while 29 are novel depression loci. For the ANX factor, 11,163 SNPs reached genome-wide significance, tagging 102 independent SNPs in 73 loci (see Figure 2a and Supplementary Table 7). Two loci overlapped with a previous GWAS of anxiety disorders (Purves et al. 2019), and 71 loci are novel for anxiety. LD Score Regression analyses indicated that our GWAS results were not due to uncontrolled inflation for either DEP (intercept = 0.991, s.e. = 0.010) or ANX (intercept = 1.003, s.e. = 0.011). Effect sizes of independent significant SNPs showed large concordance between DEP and ANX. Three variants had significantly (*P* < 2.76 × 10^−4^) different effect sizes: rs613872 was associated only with DEP, and two SNPs (rs62250713 and rs391957) with ANX only (see Figure 2b).

**Figure 2:**
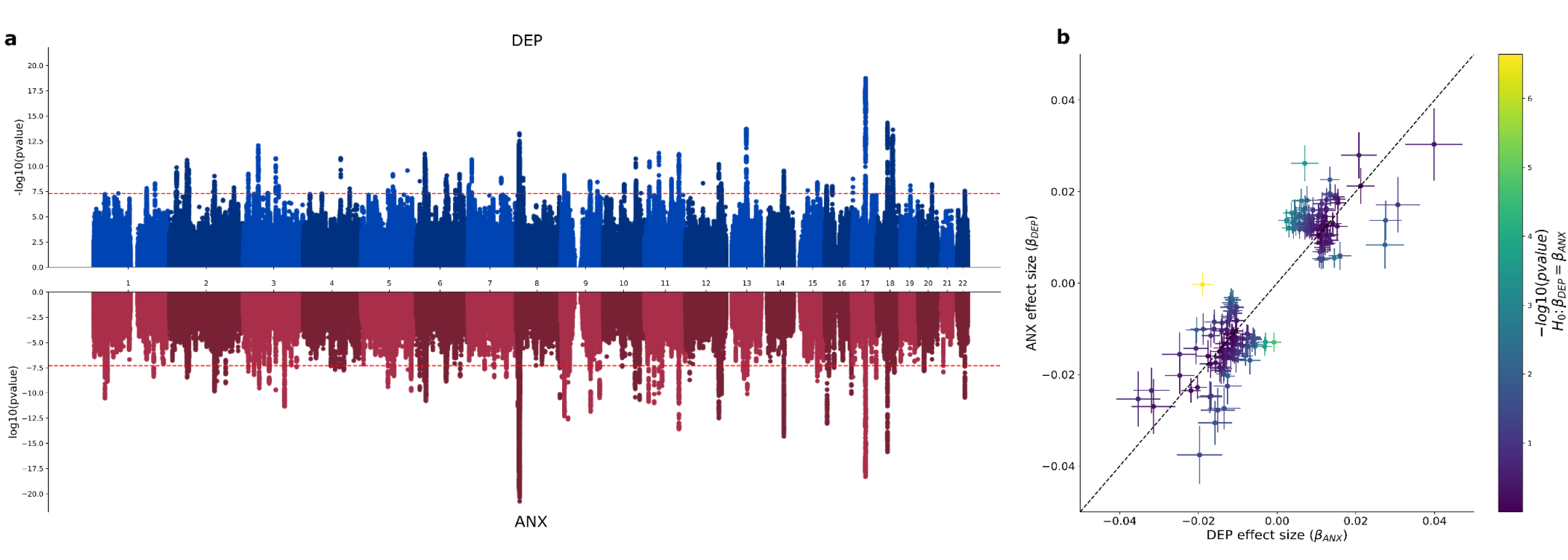
SNP-based associations of the DEP and ANX latent factors and comparison of effect sizes Note: (a) Manhattan plots for GWAS of the DEP (top; blue) and ANX (bottom; red) latent factors. The red line denotes the p-value threshold for genome-wide significance(P < 5 × 10^−8^). (b) Comparison of effect sizes for independent SNPs that reached genome-wide significance in the GWAS of either factor.

We conducted a replication of the significant independent SNPs in a cohort of research participants from 23andMe with information on self-reported diagnosis of depression (634,037 cases; 1,308,690 controls) and anxiety (624,615 cases; 1,310,854 controls). For the DEP replication, 81 variants were tested (8 SNPs were unavailable or of insufficient quality in the 23andMe cohort). Of these variants, 58 were significant after Bonferroni correction (α = 0.05/81; *P* < 6.17 × 10^−4^) and had the same direction of effect, and 40 reached genome-wide significance (*P* < 5 × 10^−8^). 93 ANX variants were tested for replication (9 SNPs were of insufficient quality in the 23andMe cohort), of which 73 were significant after Bonferroni correction (α = 0.05/93; *P* < 5.38 × 10^−4^) and had the same direction of effect, and 39 reached genome-wide significance (*P* < 5 × 10^−8^).

MAGMA was used to conduct gene-based association tests and gene-set enrichment analyses. We identified 255 genes and 9 gene sets associated with DEP, and 325 genes and 21 gene sets associated with ANX (significant after Bonferroni correction; Supplementary Tables 8-11). There was substantial overlap with respect to enriched functional categories between the two traits (110 genes and 5 gene sets).

### Polygenic risk prediction

To further validate the latent factors, we used polygenic risk scores (PRS) derived from the ANX and DEP summary statistics to predict both depressive and anxiety symptoms in an independent sample (N = 4,434). The PRS includes all SNPs, using Bayesian multiple regression to account for linkage disequilibrium (see Methods). PRS for DEP significantly predicted depressive symptoms (*P* = 2.69 × 10^−10^), explaining 1.05% of variance (see Figure 3). Similarly, PRS for ANX significantly predicted anxiety symptoms (*P* = 4.80 × 10^−14^), explaining 1.53% of variance. For comparison, we also calculated PRS from PHQ-9 sum-score, GAD-7 sum-score, neuroticism, major depression (Wray et al. 2018), depression (Howard et al. 2019) and anxiety disorders (Purves et al. 2019). The DEP and ANX latent factors explained a greater amount of variance than the sum-scores (proportional increase in explained variance was 185% and 237%, respectively), indicating that polygenic prediction was improved by the combination of leveraging information from the neuroticism items and taking a more psychometrically informed approach (i.e., factor analysis) to phenotype construction. Overall, specificity in polygenic prediction was low; PRS for depression phenotypes explained an equal or greater amount of variance in anxiety symptoms than depressive symptoms (see Figure 3 and Supplementary Table 12).

**Figure 3:**
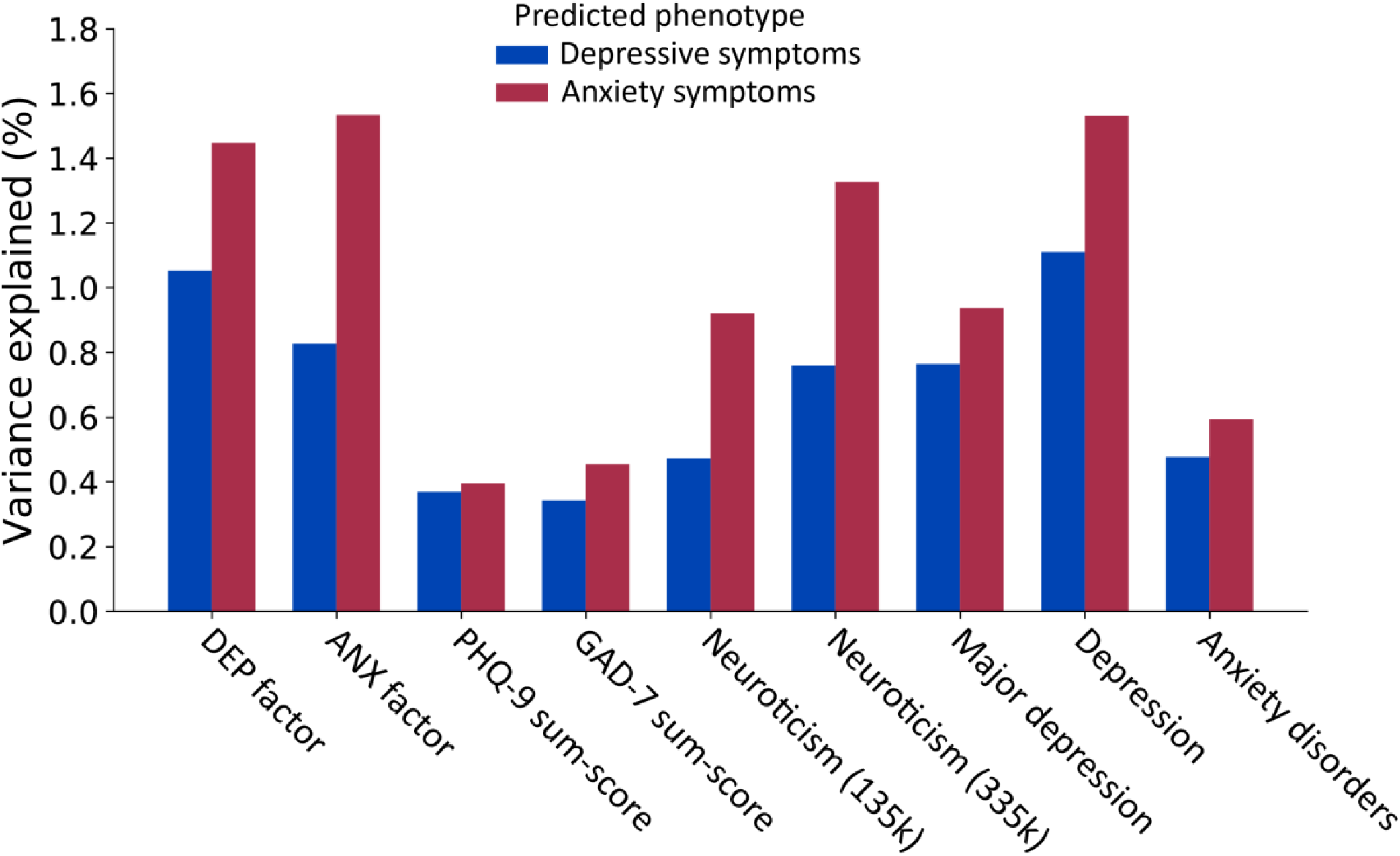
Polygenic risk prediction of depressive and anxiety symptoms Note: amount of variance in depressive and anxiety symptoms explained by PRS for the DEP and ANX latent factors, and a range of related phenotypes: PHQ-9 sum-score (N = 135,149), GAD-7 sum-score (N = 135,747), neuroticism (N = 136,212; N = 338,812), major depression (Wray et al. 2018; QIMR and 23andMe cohorts excluded; N = 159,598), depression (Howard et al. 2019; QIMR and 23andMe cohorts excluded; N = 494,258), and anxiety disorders (N = 114,019). All PRS predictions were significant after Bonferroni correction (α = 0.05/18; *P* < 2.78 × 10^−3^).

### Genetic correlations with other complex traits

We estimated genetic correlations between the ANX and DEP latent factors and a range of human complex traits (see Figure 4 and Supplementary Table 13). While patterns of correlations were similar in magnitude and direction across most of the traits, some traits showed differential genetic overlap with DEP and ANX. Smoking related phenotypes (initiation, age of initiation, cigarettes per day, and cessation) genetically correlated with DEP (|r_g_| > 0.27), but not with ANX. Genetic overlap with socioeconomic traits (Townsend deprivation index, household income, and educational attainment) was consistently larger for DEP than ANX. Conversely, ANX showed stronger overlap with obsessive compulsive disorder, anorexia nervosa, and schizophrenia.

**Figure 4:**
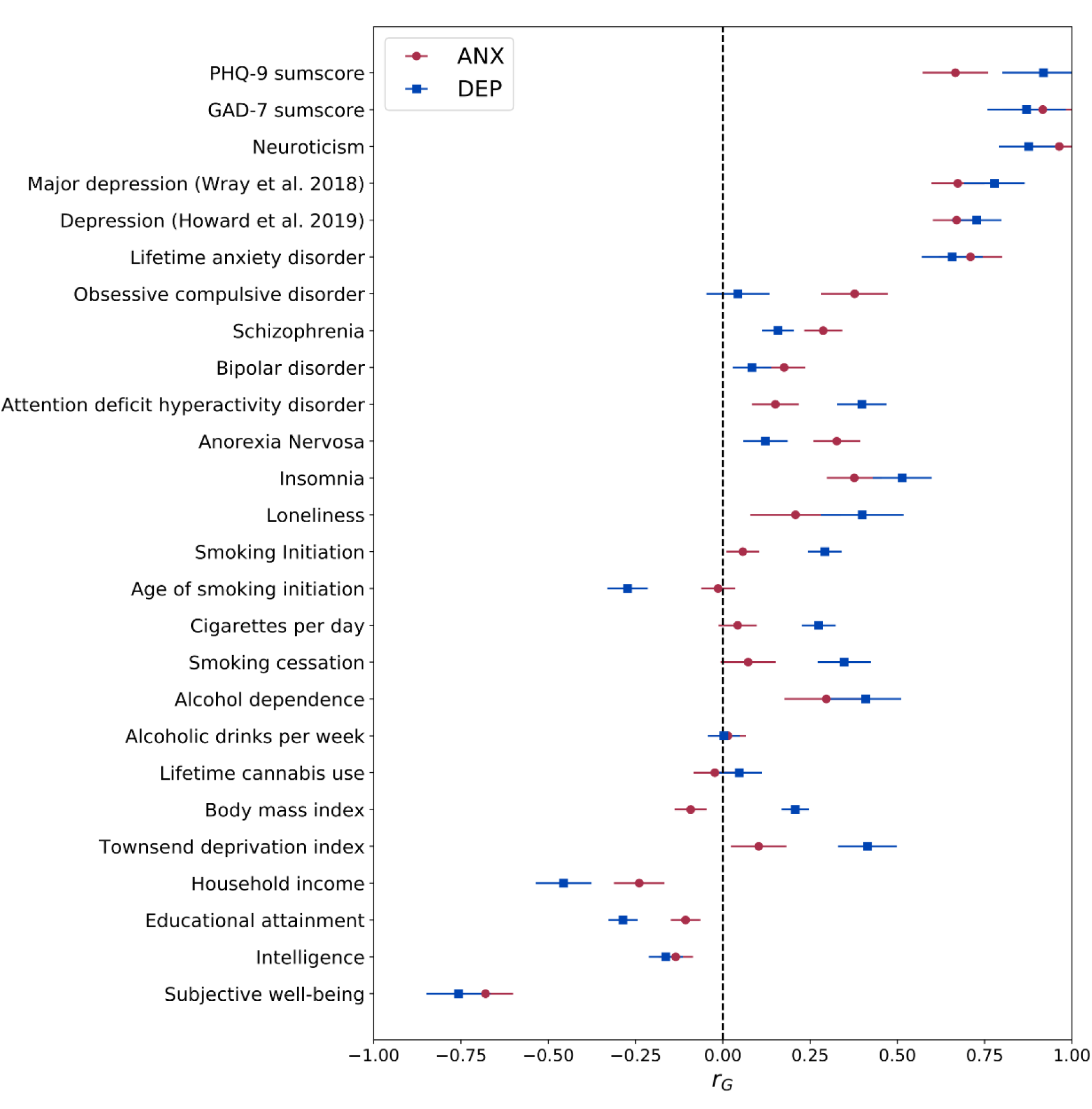
Genetic correlations with other complex traits Note: Genetic correlations between the latent factors and 26 psychiatric, substance use and socioeconomic complex traits. Error bars represent 95% confidence intervals

### Shared and trait-specific genetic influences

We sought to identify trait-specific genomic regions by conducting a pairwise analysis of the DEP and ANX GWAS summary statistics in order to characterise regions as pleiotropic, or uniquely associated with either DEP or ANX. We used gwas-pw (Pickrell et al. 2016) to estimate the posterior probability that a given genomic region is associated with (1) DEP only, (2) ANX only, (3) both DEP and ANX, and (4) both DEP and ANX but via separate causal variants. Out of the 1703 tested regions, 123 (7%) had a posterior probability greater than 0.5 of containing a causal variant for at least one of the two traits. Of these regions, 10 were uniquely associated with DEP, 20 were uniquely associated with ANX, 71 were associated with both DEP and ANX, and 22 were associated with both traits but via separate variants (see Figure 5a and Supplementary Table 14).

**Figure 5:**
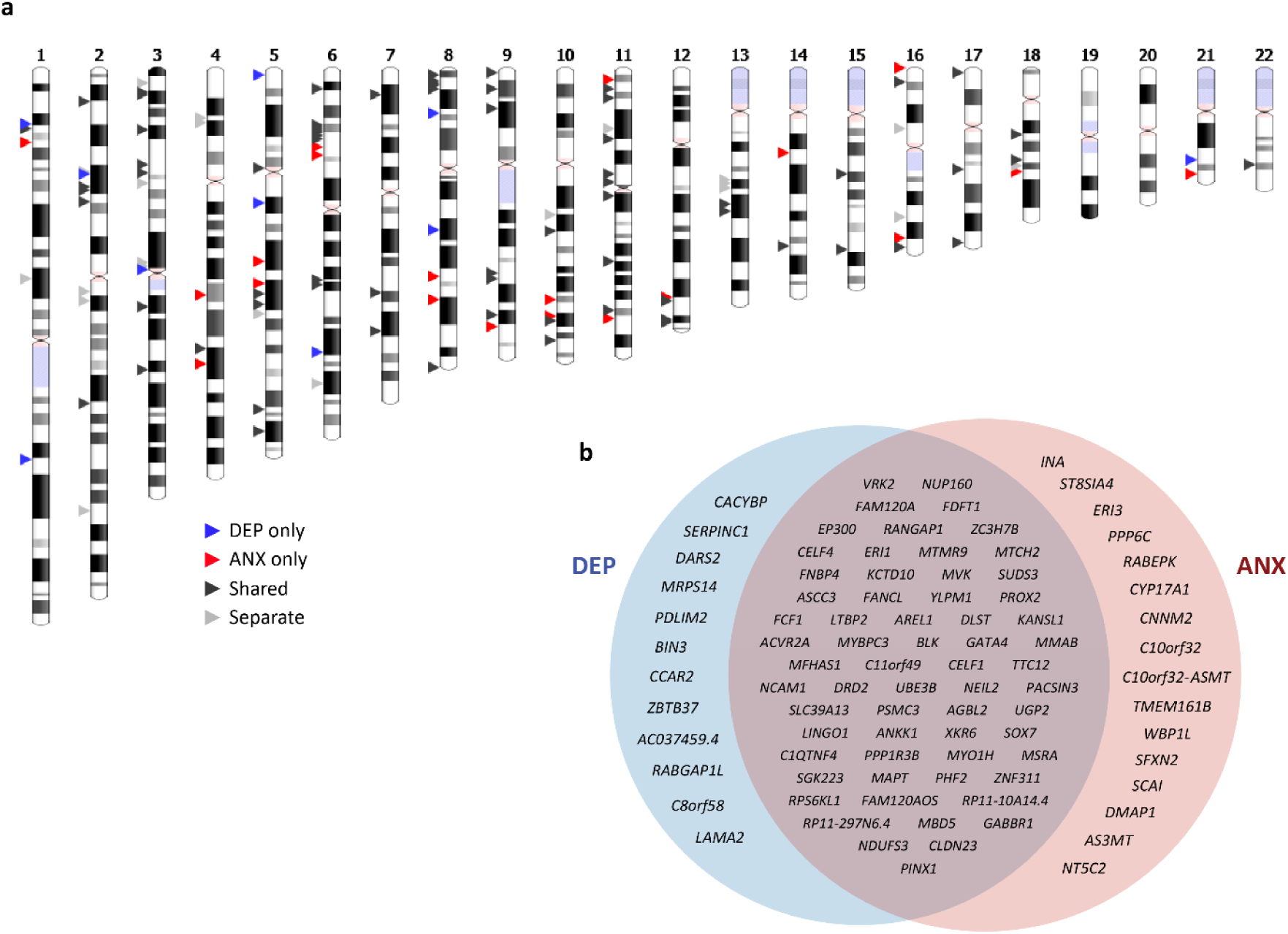
Shared and trait-specific genetic influences on depressive and anxiety symptoms. Note: (a) Genomic regions that are specific to DEP (blue arrows), specific to ANX (red arrows), shared (dark grey arrows), or influence both traits via separate variants (light grey arrows). (b) Prioritised genes for shared and trait-specific regions. Genes that reached significance (after Bonferroni correction) in the gene-based test and were identified by at least two additional methods (position, eQTL, or chromatin interaction) are presented (see Supplementary Tables 15-20 for all mapped genes).

Next, we conducted gene-based association tests separately for regions that were specific to DEP, specific to ANX, or shared. We identified 26 genes significantly associated with DEP-specific regions, 47 genes with ANX-specific regions, and 144 with shared regions (see Supplementary Tables 15-17). To further identify genes for trait-specific and shared regions we also mapped SNPs (that reached genome-wide significance in the GWAS) to genes based on proximity, eQTL, and chromatin interactions. These three strategies mapped 49 genes to DEP-specific regions, 74 genes to ANX-specific regions and 470 genes to shared regions (see Supplementary Tables 18-20). The total number of genes identified (across all methods) was 63, 102 and 509 genes for DEP-specific, ANX-specific and shared regions, respectively (see Figure 5b).

Using all genes linked to trait-specific and shared regions, we conducted gene-set enrichment analysis against gene-sets defined by traits in the NHGRI-EBI GWAS Catalog (Buniello et al. 2019). Genes prioritised for DEP-specific regions were significantly enriched in a gene-set for hypertriglyceridemia (*P* = 1.92 × 10^−7^). Genes mapped to ANX-specific regions showed enrichment in multiple gene-sets: schizophrenia (*P* = 1.99 × 10^−11^), autism spectrum disorder or schizophrenia (*P* = 2.03 × 10^−22^), response to cognitive-behavioural therapy in major depressive disorder (*P* = 1.30 × 10^−13^), and multiple gene sets related to blood pressure: mean arterial pressure (*P* = 4.37 × 10^−7^), systolic blood pressure (*P* = 3.24 × 10^−7^), pulse pressure (*P* = 2.65 × 10^−7^), and hypertension (*P* = 1.27 × 10^−5^). Genes prioritised for shared regions were significantly enriched in 109 gene-sets (see Supplementary Table 21), including autism spectrum disorder or schizophrenia (*P* = 1.71 × 10^−137^), blood protein levels (*P* = 1.04 × 10^−40^), sarcoidosis (*P* = 7.89 × 10^−33^), systolic blood pressure (*P* = 3.24 × 10^−4^), and triglyceride levels (*P* = 1.81 × 10^−3^).

## Discussion

Recent studies have revealed substantial genetic correlations between depression, anxiety, and neuroticism (Wray et al. 2018; Levey et al. 2020; Howard et al. 2019; Nagel, Jansen, et al. 2018). The extensive genetic overlap partly reflects the overlap in items used to measure these traits, which motivated us to explore the genetic factor structure underlying 28 symptoms of depression, anxiety, and neuroticism. Leveraging the underlying factor structure, we conduct GWAS on latent factors of anxiety and depressive symptoms using data from over 400,000 individuals in the UK Biobank. We identify novel and robust genetic associations and present findings for the largest GWAS of anxiety to date (102 GWS independent variants, ∼78% replicate in an independent cohort of self-reported diagnosis of anxiety). We also characterise shared and trait-specific genetic influences and report on gene-set analyses targeted at understanding shared and trait-specific aetiology.

Modelling the genetic factor structure of symptoms of depression, anxiety, and neuroticism revealed two key findings. First, symptoms of anxiety and depression loaded onto different factors, although the genetic correlation between the factors was high. This implies that while symptoms of depression and anxiety are closely related, symptoms are genetically more similar to symptoms within the same disorder than symptoms between disorders. This observation is consistent with a phenotypic-level network analysis of the PHQ-9 and GAD-7 items within a psychiatric sample (Beard et al. 2016), which found that symptom connections were higher within each disorder than between disorders. The large degree of genetic overlap between symptoms of anxiety and depression is in agreement with twin-based symptom-level analyses (Kendler et al. 1987), although our results suggest some specificity of common genetic risk factors. Second, neuroticism items loaded highly onto either the DEP or ANX factors rather than forming their own factor, suggesting that at a genetic level neuroticism is not itself a distinct construct, and likely encapsulates (sub-clinical) symptoms of both depression and anxiety. The partitioning of neuroticism items among two distinct factors is in line with the results of a hierarchical clustering analysis of the genetic correlation matrix derived from these neuroticism items when analysed in isolation (Nagel, Watanabe, et al. 2018).

By leveraging item-level genetic overlap with neuroticism, we substantially increased statistical power to identify genomic risk loci for depressive and anxiety symptoms. Our GWAS of depressive symptoms identified 62 genomic risk loci, of which 29 are novel. For anxiety symptoms, we identified 73 genomic loci (71 novel), a substantial increase from previous studies which have found 6 loci for anxiety disorders (Purves et al. 2019; Meier et al. 2019) and 6 loci for anxiety symptoms (Levey et al. 2020). The identification of a large number of replicable genomic loci for anxiety unlocks the possibility of leveraging statistical genetic approaches (e.g. Mendelian randomisation or drug repositioning) which were not possible with previous anxiety GWAS. We implemented one such approach, a regional pairwise analysis, in order to disentangle the shared genetic architecture of anxiety and depression and identify trait-specific genetic loci.

Our results suggest that depression-specific genomic regions are linked to hypertriglyceridemia. This is consistent with several studies that have found an association between depression and triglyceride levels (Igna, Julkunen, and Vanhanen 2011; Richter, Juckel, and Assion 2010; Akbaraly et al. 2009; Glueck et al. 1993), and previous GWAS of depression have reported significant genetic correlations with triglycerides (rg = 0.14; Wray et al. 2018; Howard et al. 2019). Shared regions were also enriched in a gene-set for triglyceride levels, suggesting that while both depression and anxiety contain a metabolic component, this component may be larger in depression than anxiety. Conversely, genes mapped to anxiety-specific regions were enriched in gene-sets related to multiple blood pressure phenotypes. While perhaps unsurprising given increased blood pressure is a direct physiological effect of the stress response, anxiety has also been linked to increased risk of hypertension (Pan et al. 2015). Interestingly, genes unique to ANX were also enriched in a gene-set linked to response to cognitive-behavioural therapy in major depression (Rayner et al. 2019), suggesting that the presence of comorbid anxiety symptoms may influence treatment response for depression. Indeed, comorbid anxiety and major depressive disorders are associated with higher symptom severity and impairment, disorder persistence, and reduced response rates (Young, Mufson, and Davies 2006; Kessler et al. 2007; Emmanuel, Simmonds, and Tyrer 1998; Walker et al. 2000; Altamura et al. 2004). There was also significant enrichment of ANX-specific regions in schizophrenia gene-sets, consistent with a larger genetic correlation of schizophrenia with the ANX factor than with DEP factor. Anxiety symptoms are highly prevalent in patients with schizophrenia (Achim et al. 2009) and are associated with the positive symptom domain of schizophrenia (Emsley et al. 1999).

Genetic correlations with other complex traits were largely concordant in direction and magnitude across DEP and ANX. Smoking related phenotypes however, were genetically correlated with depression but not with anxiety. Observational studies on the association of smoking with anxiety and depression are largely mixed with regards to the direction of effect (Fluharty et al. 2017). We find moderate genetic correlations between DEP and smoking initiation, cigarettes per day, cessation (positive), and age of initiation (negative). Genetic correlations were not significant between any smoking phenotype and ANX, suggesting a unique relationship between smoking behaviour and depression.

It is well established that depression and anxiety share a substantial amount of genetic liability. Our results provide additional evidence for this notion, with a high genetic correlation between the DEP and ANX factors (rg = 0.80), consistent with the genetic overlap between major depression and anxiety disorders in previous studies (rg = 0.75-0.80). Further, the amount of polygenic overlap (i.e., the fraction of genetic variants causally influencing both traits) was considerable, with 71 out of 123 genomic regions containing a causal variant that influences both traits. We note that this is likely an underestimate of the proportion of shared genetic effects, as the correction for sample overlap in pairwise GWAS is conservative and some truly shared genetic effects may be corrected out (Pickrell et al. 2016). Given the high genetic overlap and substantial comorbidity between depression and anxiety (Kessler et al. 2007; Lamers et al. 2011), it is unsurprising that there was very little differentiation in polygenic prediction of depressive and anxiety symptoms with PRS derived from latent factors. This was not specific to the latent factors; poor specificity was also seen with PRS derived from clinical depression and anxiety phenotypes (Wray et al. 2018; Purves et al. 2019).

We performed multiple analyses to validate the latent factors representing depression and anxiety. First, genetic correlations between the latent factors and the PHQ-9 and GAD-7 sum-scores (i.e., between the DEP factor and the sum-score of the 9 depressive symptoms, and between the ANX factor and the sum-score of the 7 anxiety symptoms) were very high and not significantly different from one. These correlations persisted when incorporating an additional ∼270,000 individuals with neuroticism information only into the latent factors. Second, a substantial proportion (∼72% for DEP and ∼78% for ANX) of genome-wide significant variants replicated in a large, independent cohort of self-reported diagnosis of anxiety and depression. Third, polygenic risk score analyses showed that the latent factors significantly predicted depressive and anxiety symptoms in a second independent sample.

The findings of the present study should be interpreted in light of some key limitations. The DEP and ANX latent factors represent depressive and anxiety symptoms within a population-based cohort. While these factors had relatively high genetic correlations with clinical phenotypes, it is likely that they do not capture the entire spectrum of genetic influences on major depression and anxiety disorders (Schwabe et al. 2019; Kendler et al. 2019; Cai et al. 2020). For example, the episodic nature of major depression or the persistence of generalised anxiety are not well captured by symptom questionnaires. Second, by leveraging neuroticism items in the extended UKB sample, we increased statistical power to identify variants for symptoms which overlap with depression and anxiety (predominately ‘psychological’ symptoms). However, the somatic, motor, and neurovegetative symptoms characteristic of major depression and anxiety disorders are not well represented in neuroticism, and consequently not within our phenotypes.

Over a decade of molecular genetic studies have confirmed the presence of widespread pleiotropy across psychiatric disorders (Smoller et al. 2019; Lee et al. 2019; Watanabe et al. 2019). The substantial sharing of genetic risk factors challenges the utility of analysing discrete diagnostic categories of psychopathology defined by current classification systems (such as the Diagnostic and Statistical Manual of Mental Disorders 5th edition; DSM-5) in the discovery of genetic risk loci and prediction of disease. There has been recent interest in analysing psychiatric endophenotypes as an alternative (Sanchez-Roige and Palmer 2020), with initiatives such as the Hierarchical Taxonomy of Psychopathology (HiTOP) (Waszczuk et al. 2020) and Research Domain Criteria (RDoC) (Insel et al. 2010) facilitating such efforts. Symptom-level analyses are one such endophenotypic approach which may prove useful in in advancing our understanding of the genetic aetiology of psychopathology, by allowing the discovery of symptom-specific genetic influences and the identification of genetic subtypes and trans-diagnostic factors of genetic liability. We show that analysing genetically homogenous combinations of symptoms across traits can increase statistical power to identify loci and elucidate comorbidity and genetic overlap between psychiatric phenotypes.

## Methods

### UK Biobank

Data for the main analyses came from the UK Biobank (UKB), a major health data resource containing phenotypic information on a wide range of health-related measures and characteristics in over 500,000 participants from the United Kingdom general population (Bycroft et al. 2018). Participants were excluded from the present study based on ancestry, relatedness and withdrawn consent. Participants were included if they were of white British ancestry, identified through self-reported ethnicity and ancestral principal components. Participants who self-reported as not white British, but for whom the first two genetic principal components indicated them to be genetically similar to those of white British ancestry were also included in order to maximise sample size (MacGregor et al. 2018).

### Depressive and anxiety symptoms

Depressive symptoms were assessed with the 9-item Patient Health Questionnaire (PHQ-9) (Kroenke, Spitzer, and Williams 2001), and anxiety symptoms with the 7-item Generalised Anxiety Disorder scale (GAD-7) (Spitzer et al. 2006). Over 150,000 participants completed the PHQ-9 and GAD-7 as part of a UKB mental health follow-up questionnaire administered online in 2016 (Davis et al. 2020). Each item assesses the frequency of a particular symptom over the past two weeks, rated on a four-point ordinal scale: (0) not at all, (1) several days, (2) more than half the days, (3) nearly every day.

The ordinal scale of measurement of these items complicates interpretation of SNP-based heritability (*h*^2^ _SNP_) estimates. As *h*^2^ _SNP_ estimates are utilised in Genomic SEM, we transformed each item into a binary phenotype in order for interpretable *h*^2^ _SNP_ estimates to be produced. Items were dichotomised such that an item was considered to be endorsed if the item score was one or greater (several days, more than half the days, or nearly every day), and not endorsed if the score was zero (not at all). A cut-off score of one was used in order to maximise the number of participants who endorsed an item and hence statistical power (genetic correlations between the ordinal-scale items and binary items were all > 0.95; median r_g_ = 0.98).

### Neuroticism

Neuroticism was measured using the 12-item Eysenck Personality Questionnaire-Revised Short Form (EPQR-S) (Eysenck, Eysenck, and Barrett 1985), with each item assessed on a dichotomous scale (‘yes’ or ‘no’). The questionnaire was administered to the entire UKB cohort (∼500,000 participants).

### Genome-wide association analyses

GWAS analyses of the 28 individual items (9 depression items, 7 anxiety items, 12 neuroticism items) were conducted via logistic regression in PLINK v2.00a (Chang et al. 2015). If two individuals in the sample were related (pi-hat > 0.2) one individual was removed (preferentially from the control set if the related individuals were in both case and control sets). GWAS analyses of the PHQ-9, GAD-7, and EPQR-S sum-scores were conducted via linear regression. Analyses were limited to autosomal SNPs with high imputation quality score (INFO score ≥ 0.80) and a minor allele frequency of 1% or higher, resulting in 9,417,325 SNPs being tested for association. Age, sex, genotyping array, and 20 principal components were included as covariates.

### Factor analysis

We first explored the genetic factor structure by conducting an exploratory factor analysis (EFA) based on the genetic correlation matrix of the 28 items. The EFA was conducted using only participants who completed the UKB mental health questionnaire (N range = 132,602 – 137,461; see Supplementary Table 1 for item-specific sample sizes). As participants who completed the UKB mental health questionnaire differ significantly from the entire UKB cohort (e.g. higher educational attainment, higher socio-economic status, lower rates of smoking; Davis et al. 2020), we restricted the EFA to a subset for neuroticism to ensure these systematic differences did not bias the EFA. Cross-trait LD Score Regression was used to estimate genetic correlations between each of the 28 items. These estimates are not biased by sample overlap (Bulik-Sullivan et al. 2015). The ‘psych’ R package was used to conduct the EFA, with an ordinary least squares extraction method and oblimin rotation method. Two procedures were used to decide on the optimal number of factors to extract: (1) a minimum average partial (MAP) test (Velicer 1976) (the lowest average squared partial correlation indicates the number of factors to extract), and the eigenvalue-greater-than-one rule (Kaiser 1960) (factors with an eigenvalue above 1 are extracted).

The factor model identified using EFA (retaining factor loadings > 0.4) were subsequently carried forward in a confirmatory factor analysis (CFA) in Genomic SEM (Grotzinger et al. 2019). This was done to assess the fit of the factor model to the data while taking into account uncertainty in covariance estimates, and to allow the estimation of genetic correlations between latent factors and external traits (i.e. PHQ-9 and GAD-7 sum-scores). The default Diagonally Weighted Least Squares (DWLS) estimator was used. SNP-based heritability estimates (diagonal of the genetic covariance matrix) were converted to the liability scale, where the population prevalence of the items was estimated from the UKB sample (population prevalence = sample prevalence; see Supplementary Table 1).

### Multivariate genome-wide association analyses

GWAS of the latent factors of anxiety and depressive symptoms were conducted in Genomic SEM. All summary statistics were standardized with respect to the variance in the phenotype (i.e., STDY) using the *sumstats* function in Genomic SEM. The factor structure identified in the EFA was specified as the model. SNPs tested for association in the univariate item-level GWAS and also contained in the 1000 genomes phase 3 reference sample (with MAF > 0.01) were included, resulting in the analysis of 7,746,079 SNPs. We applied the conventional genome-wide significance threshold of *P* < 5 × 10^−8^.

The results were annotated using FUMA (Watanabe et al. 2017). Significant SNPs were clumped into blocks high in linkage disequilibrium (the non-random association of alleles at a specific locus; LD) using a threshold of r^2^ < 0.10 (correlation between allele frequencies of two SNPs). Genomic risk loci were identified by merging independent SNPs if r^2^ ≥ 0.10 and their LD blocks were physically close to each other at a distance of 1000 kb.

### 23andMe replication cohort

In the 23andMe replication analysis, case-control status was determined by self-reported depression (634,037 cases; 1,308,690 controls) or self-reported anxiety (624,615 cases; 1,310,854 controls) from samples of European ancestry (close relatives removed) in the 23andMe research cohort. The self-reported phenotype of depression was defined as cases if samples have ever been diagnosed with depression, or controls if samples have never been diagnosed with depression; the self-reported phenotype of anxiety was defined as cases if samples have ever been diagnosed with anxiety, or controls if samples have never been diagnosed with anxiety. All individuals included in the analyses provided informed consent and participated in the research online, under a protocol approved by the external AAHRPP-accredited IRB, Ethical & Independent Review Services (http://www.eandireview.com).

Association analyses were conducted by 23andMe; a logistic regression assuming an additive model for allelic effects was used with adjustment for age, sex, indicator variables to represent the genotyping platforms and the first five genotype principal components. The summary statistics were provided for independent genome-wide significant SNPs in the depression and anxiety latent factors GWAS. In the replication analysis of self-reported depression, 8 SNPs were unavailable or of insufficient quality in the replication sample, therefore 81 variants were tested for replication. In the replication analysis of self-reported anxiety, 9 SNPs were of insufficient quality or unavailable in the replication samples, therefore 93 variants were tested for replication.

### Polygenic risk prediction

The target sample consisted of an adult cohort (N = 4,434) from the Australian Twin Registry. Depressive and anxiety symptoms were assessed by the Delusions-Symptoms-States Inventory: Anxiety and Depression Scales (DSSI/sAD) (Bedford, Foulds, and Sheffield 1976), which consists of seven anxiety and seven depression items. Each item assesses the degree of distress due to a particular symptom, rated on a four-point ordinal scale: (0) none, (1) a little, (2) a lot, (3) unbearably. Additional details of the cohort and assessment procedures are reported elsewhere (Jardine, Martin, and Henderson 1984).

In total, nine polygenic risk scores (PRS) were created, using SNP weights from: DEP latent factor (UKB), ANX latent factor (UKB), PHQ-9 sum-score (UKB; N = 135,149), GAD-7 sum-score (UKB; N = 135,747), neuroticism (UKB -MHQ subset; N = 136,212), neuroticism (UKB; N = 338,812), major depression (Wray et al. 2018 -QIMR and 23andMe cohorts excluded; N = 159,598), depression (Howard et al. 2019 -QIMR and 23andMe cohorts excluded; N = 494,258), and anxiety disorders (Purves et al. 2019; N = 114,019). We used SBayesR (Lloyd-Jones et al. 2019) to account for the correlation in effect sizes arising from linkage disequilibrium (LD). Briefly, SBayesR implements Bayesian multiple regression to jointly analyse all SNPs and account for LD between SNPs. As recommended, a shrunk matrix derived from ∼3 million SNPs (MAF > 0.01) on 50k participants from the UKB was used as the LD correlation matrix. Polygenic risk scores were calculated in PLINK v1.90 (Purcell et al. 2007). For each set of PRS we estimated the proportion of variance explained by the PRS in sum-scores of both anxiety symptoms and depressive symptoms in the target sample, using genomic restricted maximum likelihood (GCTA-GREML) in order to account for relatedness within the target sample. We also estimated the amount of variance explained by PRS in log-transformed and inverse normal-transformed sum-scores of depressive and anxiety symptoms (presented in Supplementary Table 12).

### Gene-based tests and gene-set analysis

MAGMA v1.07 (de Leeuw et al. 2015) was used to conduct gene-based and gene-set analyses on the summary statistics of the DEP and ANX latent factors. The gene-based analysis tested 18,756 protein coding genes for association. A Bonferroni corrected significance threshold was applied (*P* < 2.67 × 10^−6^). The gene-set analysis tested 7,250 gene sets for association with DEP and ANX factors. A Bonferroni corrected significance threshold was applied (*P* < 6.90 × 10^−6^).

### Pairwise analysis of GWAS summary statistics

The pairwise GWAS analysis was implemented using gwas-pw (Pickrell et al. 2016). First, the genome is split into 1703 approximately independent regions (Berisa and Pickrell 2016). Then the posterior probability of each of the following models is calculated: (1) the region is associated with DEP only, (2) the region is associated with ANX only, (3) the region is associated with both DEP and ANX, and (4) there are separate associations for ANX and DEP within that region. To account for sample overlap across the two traits, gwas-pw requires the correlation between effect sizes in the two traits in non-associated regions. We used fgwas (Pickrell et al. 2014) to calculate the posterior probability of association (PPA) for each region with both traits. We then calculated the correlation in effect sizes for SNPs in regions with a PPA < 0.2 for both ANX and DEP. Given one of the models has a posterior probability > 0.5, we report the model with the highest posterior probability. Results are presented in the form of an ideogram, created in the Complex-Traits Genetics Virtual Lab (Cuéllar-Partida et al. 2019).

### Gene-mapping of trait-specific or shared regions

First, MAGMA was used to conduct gene-based tests separately on genomic regions reported to be associated with only DEP (120 protein-coding genes tested; Bonferroni corrected significance threshold, *P* < 4.17 × 10^−4^), regions associated with only ANX (390 protein-coding genes tested; Bonferroni corrected significance threshold, *P* < 1.57 × 10^−4^), and regions associated with both ANX and DEP (1038 protein-coding genes tested; Bonferroni corrected significance threshold, *P* < 4.82 × 10^−5^).

Three additional methods implemented in FUMA were used to map SNPs in trait-specific or shared regions (GWAS *P* < 5 × 10^−8^) to genes. (1) Positional mapping: SNPs are mapped to genes based on proximity (within a 10kb window). (2) eQTL mapping: SNPs are mapped to a gene if they have a significant (FDR < 0.05) association with the expression level of that gene. We used eQTL information from GTEx v8 (Lonsdale et al. 2013), the CommonMind Consortium (Fromer et al. 2016), and BRAINEAC (Ramasamy et al. 2014). (3) Chromatin interaction mapping: genes are mapped if there is a significant (FDR < 1 × 10^−6^) chromatin interaction between a genomic region (within a genomic risk locus) and promotor regions of genes 250 bp upstream and 500 bp downstream of the transcription start site. Hi-C sequence data was used to identify chromatin interactions from 23 human tissue and cell types (Schmitt et al. 2016).

All prioritised genes for trait-specific and shared regions were used to conduct gene-set enrichment analyses (hypergeometric test performed in FUMA) against gene-sets defined by traits in the NHGRI-EBI GWAS Catalog (Buniello et al. 2019). Multiple testing was corrected for with a Benjamini-Hochberg false discovery rate (FDR) of 0.05.

## Data Availability

Summary statistics will be made publicly available upon publication

## Acknowledgements

We thank the research participants of all cohorts for making this study possible. This work was conducted using the UK Biobank Resource (application number 25331). NGM received funding from the Australian National Health & Medical Research Council (NHMRC) to conduct surveys in the Australian Twin Registry. SM is supported by an NHMRC Fellowship.

Members of the 23andMe Research Team include: Michelle Agee, Stella Aslibekyan, Adam Auton, Robert K. Bell, Katarzyna Bryc, Sarah K. Clark, Sayantan Das, Sarah L. Elson, Kipper Fletez-Brant, Pierre Fontanillas, Nicholas A. Furlotte, Pooja M. Gandhi, Karl Heilbron, Barry Hicks, David A. Hinds, Karen E. Huber, Ethan M. Jewett, Yunxuan Jiang, Kira Kalkus, Aaron Kleinman, Keng-Han Lin, Nadia K. Litterman, Marie K. Luff, Matthew H. McIntyre, Kimberly F. McManus, Steven J. Micheletti, Sahar V. Mozaffari, Joanna L. Mountain, Priyanka Nandakumar, Elizabeth S. Noblin, Carrie A.M. Northover, Jared O’Connell, Aaron A. Petrakovitz, Steven J. Pitts, G. David Poznik, J. Fah Sathirapongsasuti, Janie F. Shelton, Chao Tian, Joyce Y. Tung, Robert J. Tunney, Vladimir Vacic, and Xin Wang.

